# Pathway-based Rare Variant Burden Analysis Identifies a Role for the Complement System in an Extreme Phenotype of Sepsis with Coagulopathy

**DOI:** 10.1101/2022.02.24.22271459

**Authors:** Pavan K. Bendapudi, Sumaiya Nazeen, Justine Ryu, Onuralp Söylemez, Betty Rouaisnel, Meaghan Colling, Bryce Pasko, Alissa Robbins, Michael Bouzinier, Lindsay Tomczak, Lindsay Collier, Sanjay Ram, Agnes Toth-Petroczy, Joel Krier, Elizabeth Fieg, Walter H. Dzik, James C. Hudspeth, Olga Pozdnyakova, Valentina Nardi, Richard Maas, Shamil Sunyaev, Julie-Aurore Losman

**Affiliations:** Division of Hemostasis and Thrombosis, Beth Israel Deaconess Medical Center, Boston, MA; Division of Hematology and Blood Transfusion Service, Massachusetts General Hospital, Boston, MA; Harvard Medical School, Boston, MA; Division of Genomic Medicine, Brigham and Women’s Hospital, Boston, MA; Department of Medical Oncology, Dana-Farber Cancer Institute, Boston, MA; Department of Pathology, University of Colorado School of Medicine; Immunology Division, Walter and Eliza Hall Institute of Medical Research, Melbourne, Australia; Division of Infectious Diseases and Immunology, University of Massachusetts Medical School, Worcester, MA; Department of Medicine, Boston Medical Center, Boston, MA; Boston University School of Medicine, Boston, MA; Department of Pathology, Brigham and Women’s Hospital, Boston, MA; Department of Pathology, Massachusetts General Hospital, Boston, MA; Division of Hematology, Brigham and Women’s Hospital, Boston, MA

## Abstract

Extreme disease phenotypes have the potential to provide key pathophysiologic insights, but the study of these conditions is challenging due to their rarity and the limited statistical power of existing methods. Herein, we apply a novel pathway-based approach to investigate the role of rare genomic variants in infectious purpura fulminans (PF)^1,2^, an extreme phenotype of sepsis with hyperinflammation and coagulopathy for which the role of inherited risk factors is currently unknown. Using whole exome sequencing, we found a significantly increased burden of rare, putatively function-altering coding variants in the complement system in patients with PF compared to unselected patients with sepsis (*p*-value = 0.01). Functional characterization of a subset of PF-associated variants in integrin complement receptors 3 and 4 (CR3 and CR4) revealed that they exhibit a pro-inflammatory phenotype. Our results suggest that rare inherited defects in the complement system predispose individuals to the maladaptive hyperinflammatory response that characterizes severe sepsis.

---

Rare coding variants are instrumental in identifying genes and pathways involved in disease biology. However, analyses of rare variants using traditional single-variant and single-gene association tests often fail even with large sample sizes^3^, and the assembly of patient cohorts with uncommon diseases presents an additional challenge. Two strategies can overcome these barriers: (i) conducting extreme phenotype studies (EPS) to amplify the effects of rare variants^21-24^, and (ii) leveraging prior biological knowledge by applying collapsing tests at the level of pathways in which multiple genes function together with directionally consistent effects^4-6^.

In sepsis, dysregulation of the host response to infection causes harmful levels of systemic inflammation, leading to end-organ damage^7^. The clinical acuity of sepsis varies dramatically, and it is currently impossible to predict which patients will experience a severe disease course. Attempts to identify genetic factors underlying the risk for sepsis have been limited to small genome wide association studies (GWAS)^8-11^ and a single whole exome sequencing study of 74 patients^12^. However, due to the heterogeneity of the patient population, the lack of a narrowly defined clinical phenotype, the diversity of pathogens involved in sepsis, and the inherent limitations of GWAS, these efforts have been largely inconclusive. A well-designed EPS focusing on pathways that could plausibly contribute to disease pathophysiology has the potential to overcome these challenges and elucidate novel biology that is generalizable to the broader population of septic patients.

To identify genetic risk factors for severe sepsis, we undertook a case-controlled EPS of the complement system in infectious PF, an exceptionally rare and devastating extreme sepsis phenotype^1^. The defining feature of PF is uncontrolled systemic inflammation and thrombotic coagulopathy resulting in microvascular thrombosis, end-organ damage, and death. Mortality is as high as 80%, and survivors typically experience extensive scarring and dismemberment^1,2,13^. The complement system (**Figure 1A**), which is responsible for the initial inflammatory response to infection, has long been considered a key driver of inflammation in severe sepsis and PF^14-17^. Microbes directly activate the humoral complement cascade, a tightly-regulated series of proteolytic reactions that result in pathogen destruction by the membrane attack complex (MAC) and release of highly inflammatory microbial cell components. Activated complement proteins also function as potent inflammatory mediators (anaphylatoxins) that promote shock and other features of sepsis. During this process, the complement fragment C3b is deposited on microbial surfaces, where it is converted to iC3b, a potent opsonin that binds integrin complement receptors CR3 and CR4 on macrophages^18,19^. This leads to phagocytosis of microbes and activation of immunomodulatory signaling. Integrins are each composed of an α and a β subunit, different combinations of which give rise to receptors with distinct physiologic roles. CR3 (αMβ2) is composed of integrin β2 (*ITGB2*) paired with integrin αM (*ITGAM*), whereas CR4 (αXβ2) is composed of integrin β2 paired with integrin αX (*ITGAX*). Intracellular signaling via CR3 promotes an anti-inflammatory and immunoregulatory cytokine profile, whereas CR4 signaling promotes a pro-inflammatory state (**Figure 1B**)^20-22^. Given that inflammation stemming from infection is strongly associated with aberrant activation of coagulation^23-25^, we reasoned that rare function-altering mutations in the complement system could plausibly contribute to the pathophysiology of PF.

**Figure 1:**
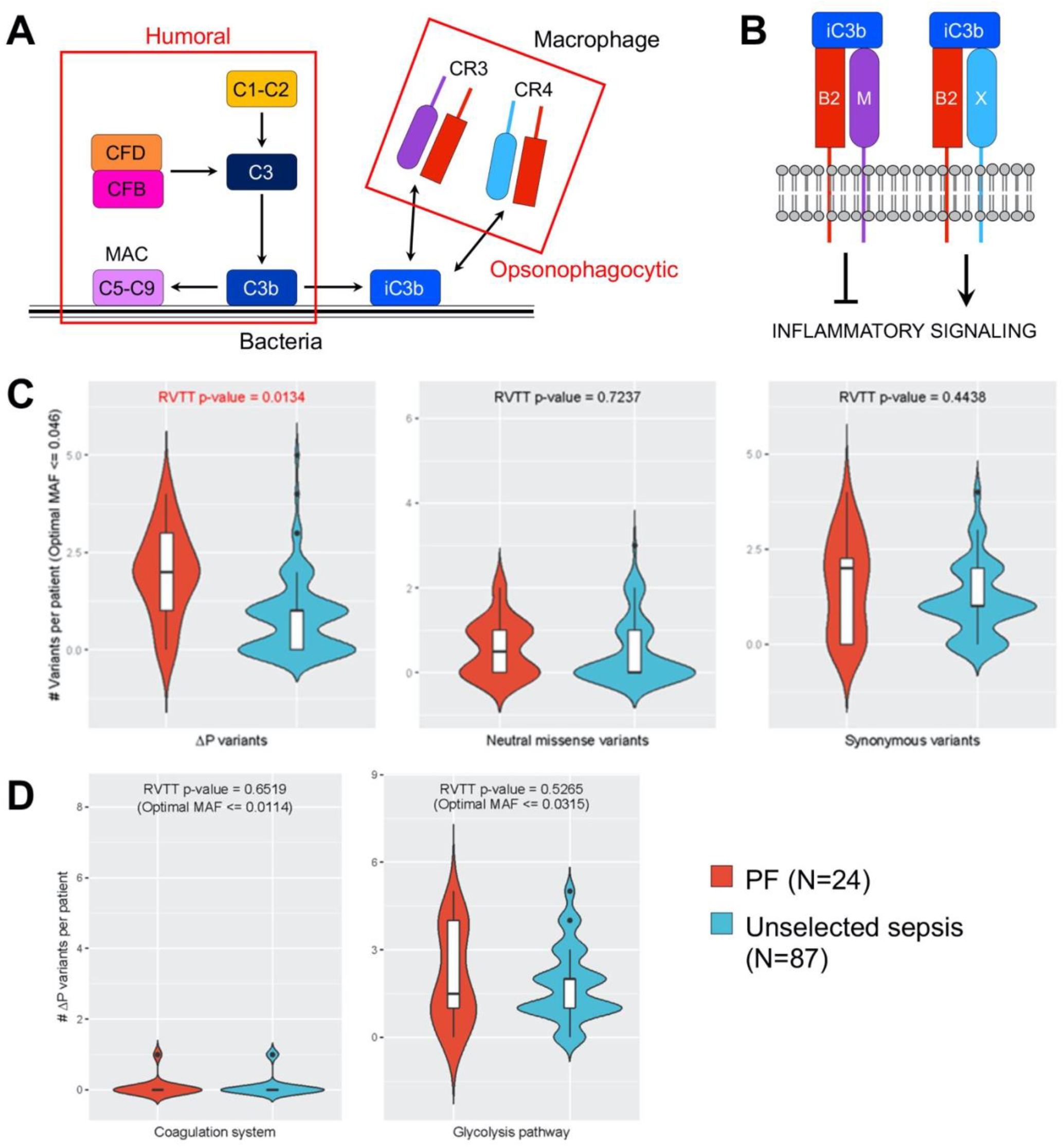
Mutational Analysis of Purpura Fulminans. (**A**) Schematic of the complement system. The complement system is broadly divided into two sub-pathways: humoral and opsonophagocytic. The humoral sub-pathway is comprised of soluble plasma proteins responsible for destroying microorganisms via formation of the membrane attack complex (MAC), which permeabilizes the surface of the target cell. The opsonophagocytic sub-pathway is comprised of cellular receptors that recognize complement fragments deposited on microbial surfaces and facilitate phagocytosis and pathogen clearance. (**B**) Heterodimerization of integrin β2 (B2) with integrin αM (M) forms the anti-inflammatory signaling mediator complement receptor 3 (CR3) whereas heterodimerization of B2 with integrin αX (X) forms the pro-inflammatory signaling mediator complement receptor 4 (CR4). (**C**) RVTT analyses comparing the European PF Cohort (PF) and the NHLBI ARDSnet iSPAAR cohort (Unselected Sepsis) in terms of ΔP variants (left panel), neutral missense variants (middle panel), and synonymous various (right panel). (**D**) RVTT analyses comparing the two cohorts in terms of ΔP variants in the coagulation system (left panel) and glycolysis pathway (right panel).

To assemble a sequencing cohort of patients with this rare, highly lethal condition, we combined prospective sample collection with a unique automated “look back” approach in which we leveraged natural language processing to perform enterprise-scale deep mining of the electronic medical record system to identify historical cases of PF. Using this technique, we were able to locate archived pathology specimens suitable for germline sequencing from PF patients who were deceased or lost to follow up. In total, we identified 40 clinically well-annotated PF patient samples and, as controls, utilized 87 unselected patients with sepsis from the NHLBI ARDSnet iSPAAR consortium (**Supplementary Tables 1 and 2**). Whole exome sequencing data from all 127 samples were jointly called using the GATK pipeline v3.5. After performing variant-level and sample-level quality control (QC), we removed three PF patient samples (**Supplementary Figure 1**). In the remaining specimens, we analyzed qualifying variants in the complement system with a gnomAD global minor allele frequency (MAF) of < 0.05 as well as those with an optimal in-cohort frequency cutoff selected using a variable thresholds (VT) approach^26^. Missense variants predicted to be “deleterious” or “possibly deleterious” by either PolyPhen2 or SIFT were grouped with high-precision loss of function (LoF) variants (stop gained, stop lost, splice donor, splice acceptor, splice region variant, and frameshift and in-frame indel) and together were termed “ΔP variants.”

Interrogating the effects of rare variants in a pathway using an EPS design requires addressing two issues. Firstly, population structure can inflate the false positive rate in both the common and rare variant space^27^. This issue can be mitigated by analyzing EPS cohorts of a single ancestry. Secondly, existing pathway-based collapsing approaches frequently focus only on the presence or absence of rare variants and are of limited value because many individuals will have at least one rare variant in a large pathway.

This challenge necessitates a new test that focuses on the number of qualifying rare variants rather than simply their presence or absence.

To assess population structure, we performed principal component analysis (PCA) on the study samples (**Supplementary Figure 2**). We found that all 87 sepsis controls but only 24/37 PF patients were of European descent, and we therefore limited our primary analysis to individuals of European ancestry. To rule out the possibility that other unrecognized population structures could introduce bias into our analysis, we evaluated the distribution of ultra-rare (i.e., doubletons and tripletons) missense and synonymous variants shared between the 24 European PF cases and 87 controls and observed no significant differences (**Supplementary Table 3**).

Across the entire Boston PF Cohort (N=37), we identified a total of 58 unique missense variants in the complement system (N = 27 autosomal genes) (**Supplementary Table 4**) with MAF < 0.05, of which 42 were present only in the PF cohort and 16 were present in both the PF and control cohorts (**Table 1 and Supplementary Tables 5 and 6**). Of these, 32 (55.2%) were annotated as functionally significant *in silico*. We also found 26 high-precision LoF variants. Of these, 22 were specific to PF patients and 4 were found in both PF and control patients. In total, 58 unique ΔP variants were identified in PF patients. In the control cohort of unselected patients with sepsis (N=87), we identified a total of 68 unique rare missense variants, of which 52 were present only among the controls and 16 were present in both cohorts. Of these, 39/68 (57.3%) were putatively function altering. The control cohort also contained 15 high-precision LoF variants, for a total of 54 ΔP variants. Of note, inclusion of non-European PF patients increased the burden of rare ΔP variants when compared to the European-only PF patient cohort (**Supplementary Figure 3**), which suggests that PF patients have an increased proportion of rare ΔP variants in the complement system regardless of ethnic background. However, we cannot validate this conclusion without a control cohort having the same population structure.

**Table 1:** Complement system variants found in the study cohorts. The number and types of complement system variants identified in the Boston PF cohort (PF) (N = 37) and the NHLBI ARDSnet iSPAAR Unselected Sepsis (US) cohort (N = 87).

|  | **Total PF-associated variants** | **Total US associated-variants** | **PF-specific variants** | **US-specific variants** | **Shared variants** |
| --- | --- | --- | --- | --- | --- |
| Neutral | 26 | 29 | 16 | 19 | 10 |
| Missense | 58 | 68 | 42 | 52 | 16 |
| Missense<sup>F</sup> | 32 | 39 | 26 | 33 | 6 |
| HP LoF | 26 | 15 | 22 | 11 | 4 |
| ΔP variants | 58 | 54 | 48 | 44 | 10 |

Missense<sup>F</sup> = missense variants predicted *in silico* to cause altered protein function
HP LoF = high-precision loss-of-function
ΔP variants = Missense<sup>F</sup> + HP LoF

To determine whether ΔP variants are enriched in PF patients, we devised a novel rare variant trend test (RVTT) utilizing the Cochran-Armitage statistic to assess the relationship between complement system variant burden and disease status^28,29^. We performed RVTT on the European PF patients (N=24) and sepsis controls (N=87) using a fixed gnomAD MAF cutoff of 0.05 as well as a variable thresholds approach with or without an in-cohort MAF cutoff. We observed a significant linear trend in the number of complement system ΔP variants in the PF patients compared to controls (gnomAD cutoff: permutation p-value = 0.0168, z-score = 2.5; VT approach with MAF cutoff up to 0.05: permutation p-value = 0.0134, z-score = 2.62; VT approach without cutoff: p-value = 0.0278, z-score = 2.72) (**Table 2, Figure 1C**). To determine whether the association of ΔP variants with PF was independent of clinical and demographic parameters, we generated a multivariate logistic regression model incorporating age, sex, and Sequential Organ Failure Assessment (SOFA) score, a validated composite measure of illness severity in critical care patients that encompasses 8 distinct clinical criteria^30^. After adjusting for these co-variates, we found that the number of ΔP variants per subject independently predicted PF in the patients of European ancestry (OR=2.61 per ΔP variant, 95% CI: 1.29-6.1, P=0.01) (**Table 3**). Including all patients in the analysis yielded a similar result. This finding suggests that multiple ΔP variants in a single patient contribute to the development of PF and orthogonally confirms the association found using RVTT.

**Table 2.** Rare variant burden assessed by Rare Variant Trend Test (RVTT) in quality-filtered European PF cases (N=24) and sepsis controls (N=87). We applied RVTT on three gene sets: the complement system, the coagulation system, and the glycolysis pathway. For each gene set, we tested rare ΔP, synonymous, and neutral missense variants. Qualifying rare variants were selected using three settings: (i) fixed threshold: gnomAD MAF < 0.05, (ii) variable threshold with no cutoff, and (iii) variable threshold with a cutoff of MAF = 0.05. We observed a significant enrichment of rare ΔP variants in the complement system under all three settings.

| Type | RVTT-Fixed Test Statistic (gnomAD cutoff) | RVTT-Fixed Perm p-value (gnomAD cutoff) | RVTT-Variable Selected Threshold (No Cutoff) | RVTT-Variable Test Statistic (No Cutoff) | RVTT-Variable Perm p-value (No Cutoff) | RVTT-Variable Selected Threshold (cutoff = 0.05) | RVTT-Variable Test Statistic (cutoff = 0.05) | RVTT-Variable Perm p-value (cutoff = 0.05) |
| --- | --- | --- | --- | --- | --- | --- | --- | --- |
| Complement System Pathway |  |  |  |  |  |  |  |  |
| $\Delta P$ variants | 2.4951 | **0.0168** | 0.0833 | 2.7229 | **0.0278** | 0.0463 | 2.6236 | **0.0134** |
| Synonymous | 1.5071 | 0.1368 | 0.25 | 1.1308 | 0.8166 | 0.0463 | 1.0267 | 0.4438 |
| Neutral missense | 1.0195 | 0.323 | 0.3704 | 1.0636 | 0.8954 | 0.0463 | 0.7881 | 0.7237 |
| Coagulation Pathway |  |  |  |  |  |  |  |  |
| $\Delta P$ variants | 1.3329 | 0.1866 | 0.015 | 0.9856 | 0.5718 | 0.0114 | 0.9856 | 0.6519 |
| Synonymous | 1.68 | 0.0996 | 0.205 | 1.4318 | 0.3251 | 0.0114 | 1.2291 | 0.4873 |
| Neutral missense | 0.7912 | 0.4864 | 0.068 | 1.3924 | 0.4095 | 0.0265 | 0.8997 | 0.7392 |
| Glycolysis Pathway |  |  |  |  |  |  |  |  |
| $\Delta P$ variants | 0.0536 | 0.9563 | 0.035 | 1.0827 | 0.5804 | 0.0315 | 1.0827 | 0.5265 |
| Synonymous | 0.1035 | 0.9256 | 0.185 | 1.4843 | 0.309 | 0.0079 | 0.8554 | 0.7307 |
| Neutral missense | 0.3942 | 0.7607 | 0.169 | 1.8254 | 0.2089 | 0.0118 | 0.6315 | 0.7491 |

**Table 3:**
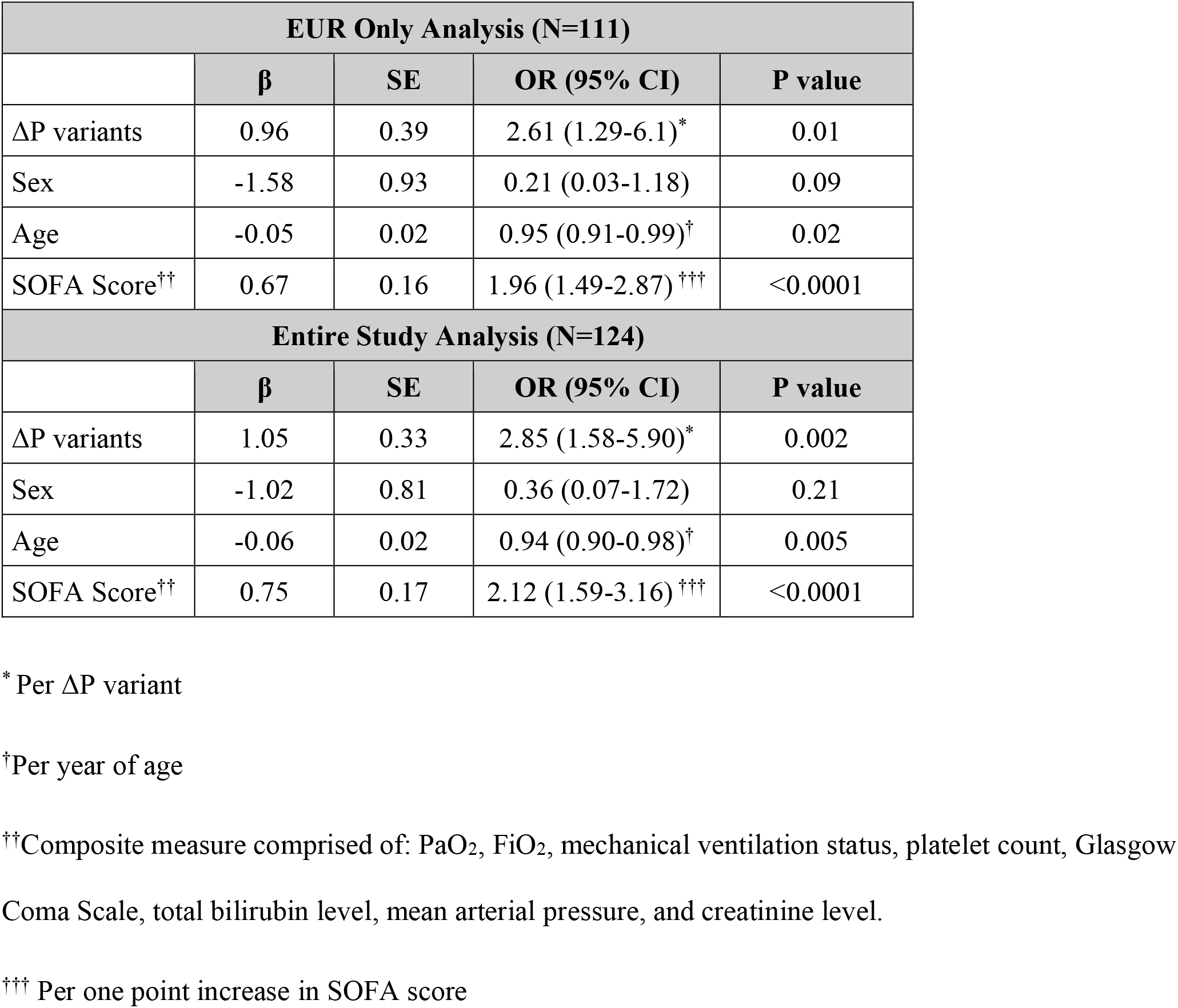
Multivariate logistic regression model of predictors of purpura fulminans.

Notably, a simple one-sided collapse and combine (CMC) Fisher’s exact test of ΔP variants in the complement system at a gnomAD global MAF cutoff of 0.05 was borderline significant (p-value: 0.0418) (**Supplementary Table 7**), and when we performed gene-based collapsing analyses using the CMC-Fisher^31^, VT-Price^26^, KBAC^32^, and SKAT^33^ tests, none of the genes in the complement system individually showed a significant enrichment of rare ΔP variants in PF cases after Bonferroni correction (**Supplementary Table 8**). This result further supports our approach of performing a pathway-based collapsing analysis that leverages prior biological knowledge. Finally, to rule out the possibility that the observed ΔP RVTT signal resulted from a technical artifact, we repeated RVTT with synonymous and predicted functionally neutral missense variants in the complement pathway and observed no significant linear trend (**Table 2, Figure 1C**). To further ensure the validity of our findings, we also performed RVTT on two additional gene sets: (i) the coagulation system, a distinct pathway that could plausibly be implicated in PF, and (ii) glycolysis, an unrelated pathway (**Supplementary Table 4**). Neither gene set demonstrated an enrichment in ΔP variants in the PF cohort (**Table 2, Figure 1D**).

Of the 58 unique ΔP variants identified in PF patients, 15 (25.9%) were found in genes encoding subunits of CR3 and CR4. Given the dearth of functional data on these variants and their high frequency in PF patients, we cloned all 11 *ITGB2, ITGAM*, and *ITGAX* variants found in the Boston PF cohort (**Supplementary Figure 4**) and characterized the resultant CR3 and CR4 receptors. We found that intracellular expression levels of all the cloned variants were similar (**Supplementary Figure 5**). However, cell-surface expression of the CR3 variants was significantly lower than that of wild-type CR3 (**Figure 2A**), whereas cell-surface expression of the CR4 variants was similar to that of wild-type CR4 (**Figure 2B**). Notably, the D283N mutation in the shared β2 subunit resulted in complete loss of CR3 surface expression without affecting the surface level of CR4. We also assessed binding of the variants to iC3b. Of the CR3 variants, 6/8 (75%) demonstrated significantly reduced binding to iC3b compared to wild-type CR3 (**Figure 2C**). By contrast, 4/7 (57.1%) CR4 variants showed similar iC3b binding to wild-type and, unexpectedly, 3/7 (42.9%) demonstrated increased binding compared to wild-type CR4 (**Figure 2D**). Given that the mutant CR4 lines all expressed equivalent or slightly reduced surface levels compared to wild-type, the observed increase in iC3b binding to specific CR4 mutants likely reflects an increased affinity of those mutants for iC3b. Finally, we used a dual-luciferase reporter assay to assess the impact of the PF-associated CR3 and CR4 variants on TNFα-mediated activation of the canonical pro-inflammatory transcription factor NF-κB. Relative to untransfected control cells, NF-κB activation was suppressed by expression of wild-type CR3 and was enhanced by expression of wild-type CR4 (**Figure 3A**). The 7 expressed CR3 variants were associated with increased activation of NF-κB compared to wild-type CR3. 6/7 (85.7%) CR4 variants showed increased NF-κB activation compared to wild-type CR4. Taken together, these findings suggest that PF-associated CR3/4 variants result in a relative loss of anti-inflammatory CR3 function and a relative gain of pro-inflammatory CR4 function.

**Figure 2:**
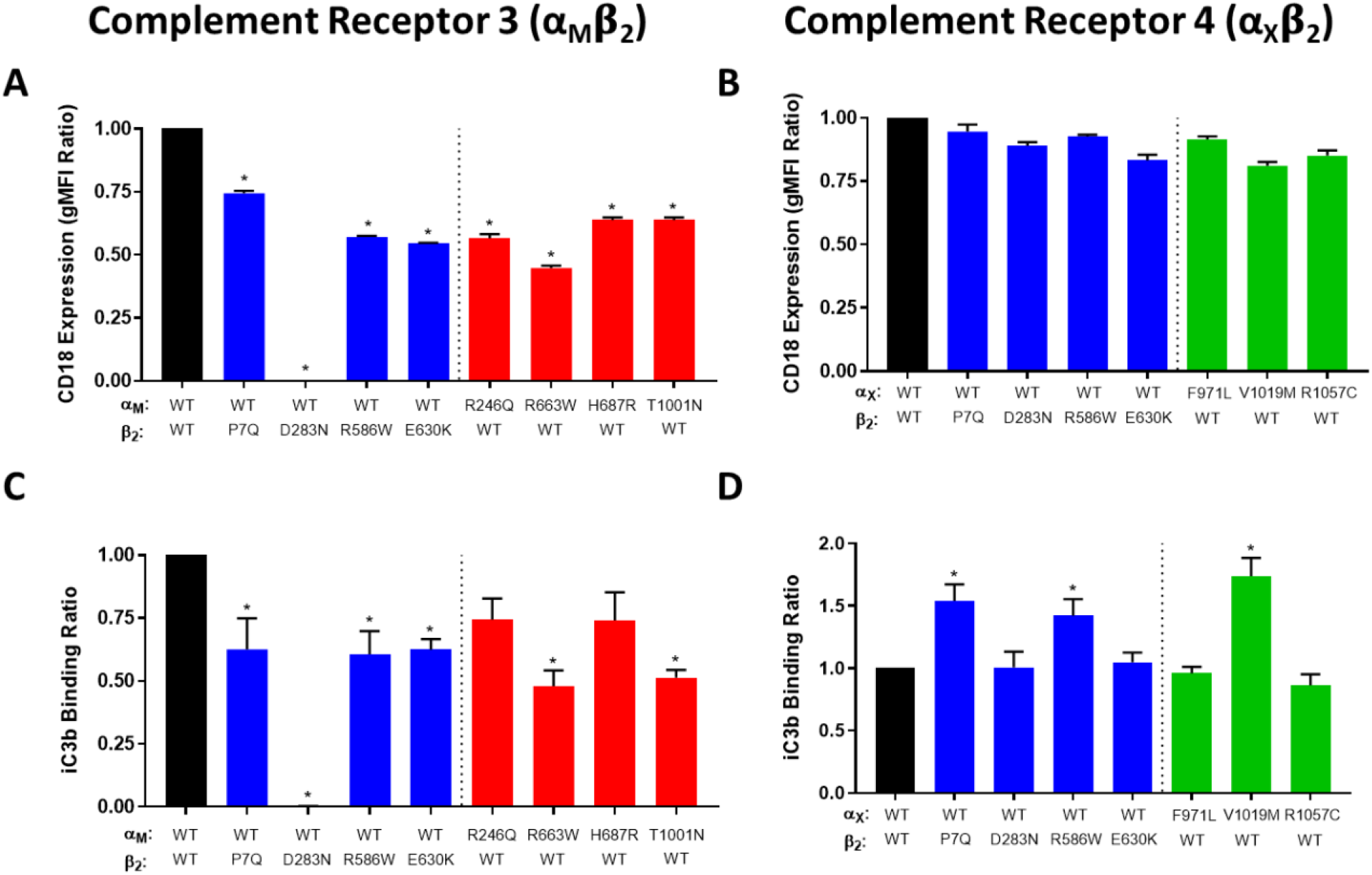
Expression and ligand binding of the CR3 and CR4 variants. (**A-B**) Wild-type (WT) and mutant complement receptors were cloned and expressed in HEK-293T cells, and surface expression levels of CR3 (A) and CR4 (B) were evaluated by flow cytometry for integrin β2 (ITGB2/CD18). WT CR3 and CR4 are depicted in black. Receptors containing a mutant integrin β2 subunit are depicted in blue, those with a mutant integrin αM (ITGAM) subunit are depicted in red, and those with a mutant integrin αX (ITGAX) subunit are depicted in green. (**C-D**) The ability of receptor-expressing cells to bind immobilized iC3b was assessed in a solid-phase functional assay. iC3b binding activity of WT and mutant CR3 (C) and WT and mutant CR4 (D) are shown. ITGB2 P7Q, ITGAM R663W, ITGAM H687R, ITGAM T1001N, and ITGAX V1019M were predicted *in silico* to be benign. ^*^P<0.05 compared to WT/WT control.

**Figure 3:**
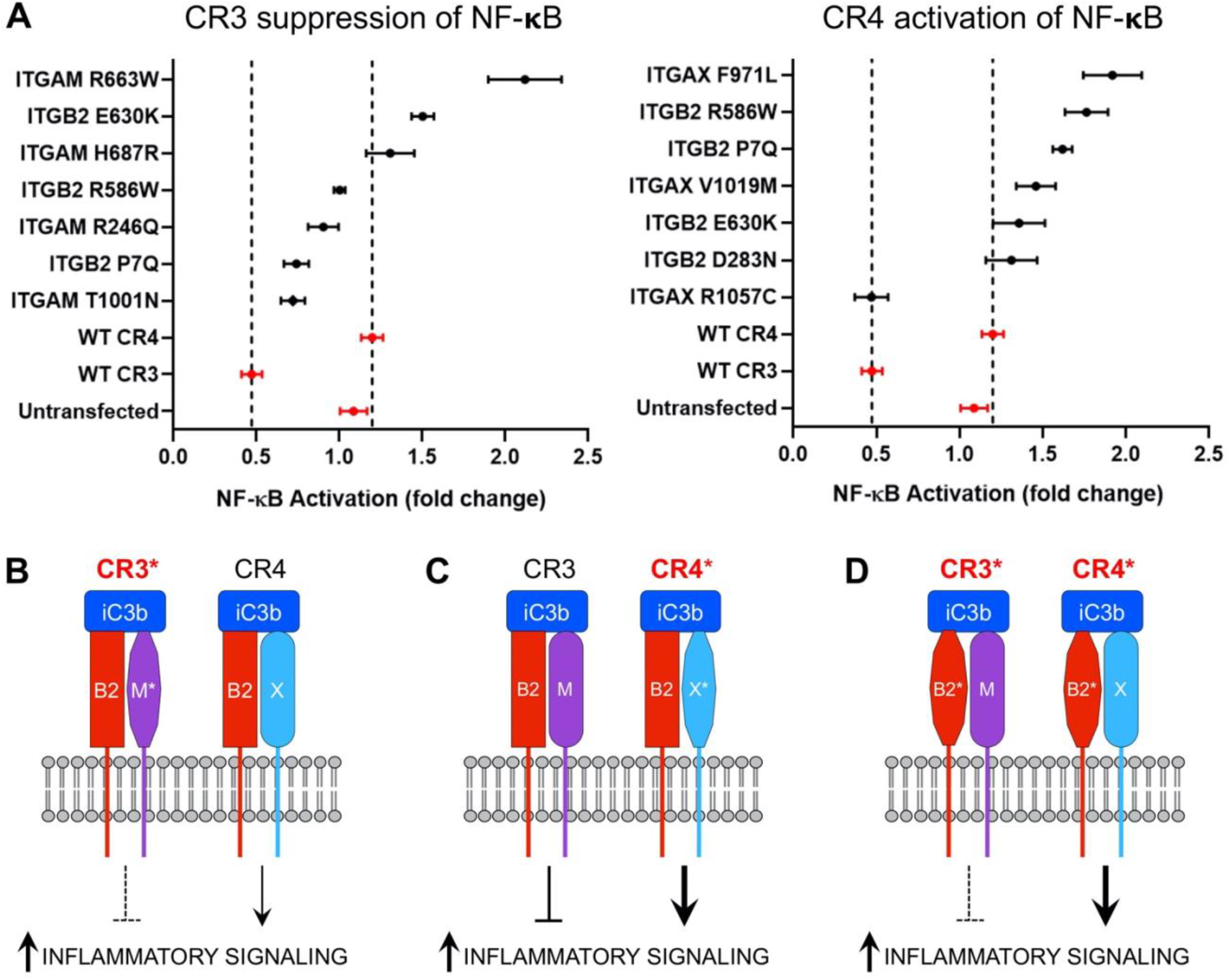
Signaling by the CR3 and CR4 variants. (**A**) TNFα-mediated activation of an NF-κB dual luciferase reporter in cells expressing the PF-associated CR3 variants (left panel) or CR4 variants (right panel) in the presence or absence of the CR3/CR4 ligand iC3b. Control HEK-293T cells not expressing CR3 or CR4 (untransfected) and HEK-293T cells expressing wild-type or mutant CR3 or CR4 were used as negative and positive controls, respectively. Data are shown as the mean fluorescence intensity ratio of the iC3b-plus condition to the iC3b-minus condition. Error bars represent standard error of the mean. (**B**) PF-associated mutations in *ITGAM* (M*) reduce the ability of mutant CR3 (CR3*) to suppress inflammatory signaling. (**C**) PF-associated mutations in *ITGAX* (X*) enhance the ability of mutant CR4 (CR4*) to activate inflammatory signaling. (**D**) PF-associated mutations in the shared subunit *ITGB2* (B2*) simultaneously decrease suppression of inflammatory signaling by CR3* and increase activation of inflammatory signaling by CR4*.

In summary, we applied RVTT, a new pathway-based mutational burden analysis approach, to an EPS of sepsis and demonstrated a relationship between rare, putatively function-altering complement system variants and the development of infectious PF. Functional characterization of PF-associated complement receptor variants identified mutations that result in loss of anti-inflammatory signaling (**Figure 3B**) or gain of pro-inflammatory signaling (**Figure 3C**), consistent with the phenotype of severe sepsis with coagulopathy^34^. We also identified several examples of single mutations in the shared complement receptor subunit integrin β2 that simultaneously enhance pro-inflammatory signaling and abrogate anti-inflammatory signaling (**Figure 3D**), a novel finding in complement receptor biology. Our genetic and functional studies support a model in which PF is driven by a genetically-encoded maladaptive inflammatory response to infection, findings that will help to risk-stratify patients with sepsis and inform the development of novel strategies to treat the thrombo-inflammatory complications of severe infection. This study also serves as a model for leveraging known biological pathways to gain insights into the genetics of extreme phenotypes and rare conditions.

Several questions remain that warrant further investigation. Firstly, due to the limited availability of clinical cases, we were unable to analyze the roles of specific microbes in the pathogenesis of PF. Secondly, our approach cannot rule out that other, as yet unidentified, genetic modifiers influence the predisposition to PF. Despite these limitations, this study represents an important step forward in defining the genetic determinants of severe sepsis.

## Supporting information

Online Methods, Supplementary Figures, Tables, and Disclosures

## Data Availability

Raw whole-exome sequencing data from PF patients will be available upon request. Raw whole-exome sequencing data from unselected sepsis patients is available from the NHLBI ARDSnet iSPAAR consortium (dbGaP Study Accession: phs000631.v1.p1). The variant call file containing the jointly-called sequence variants passing GATK filters that were analyzed in this paper will be available through dbGaP. The source code for RVTT is available through https://github.com/snz20/RVTT. Source data for all figures and tables are provided with the manuscript.

https://www.ncbi.nlm.nih.gov/projects/gap/cgi-bin/molecular.cgi?study_id=phs000631.v1.p1

