## Supplementary material for "Pathway-based Rare Variant Burden Analysis Identifies a Role for the Complement System in an Extreme Phenotype of Sepsis with Coagulopathy": Online Methods, Supplementary Figures, Tables, and Disclosures

Bendapudi, P., et al.

This PDF file includes:

Supplementary Figures (5 figures)

Supplementary Tables (8 tables)

Methods

Supplementary References

Acknowledgements

Author Contributions

Competing Interests

**Supplementary Figure 1. Per-sample Ti/Tv ratio in the dataset.** Boston PF cohort samples (N=40) are colored in blue and NHLBI ARDSnet iSPAAR control cohort samples (N=87) are colored in orange. The dotted line indicates the sample average, and the dashed lines indicate the 3 standard-deviation bounds on the sample average. Outlier samples (circled in red) were excluded from the analysis.

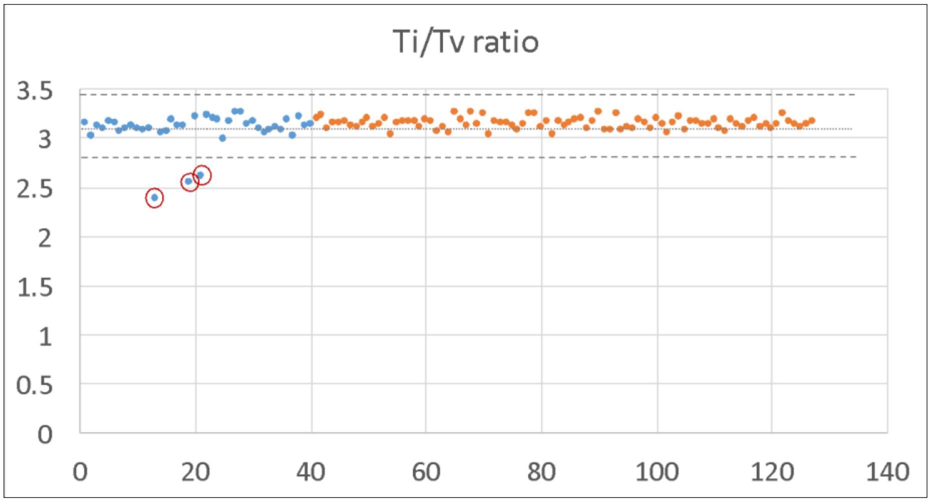

**Supplementary Figure 2: Principal component analysis of the study cohorts.** The ancestry compositions of the Boston PF cohort (PF) and the NHLBI ARDSnet iSPAAR cohort (NHLBI Sepsis), as determined by principal component analysis.

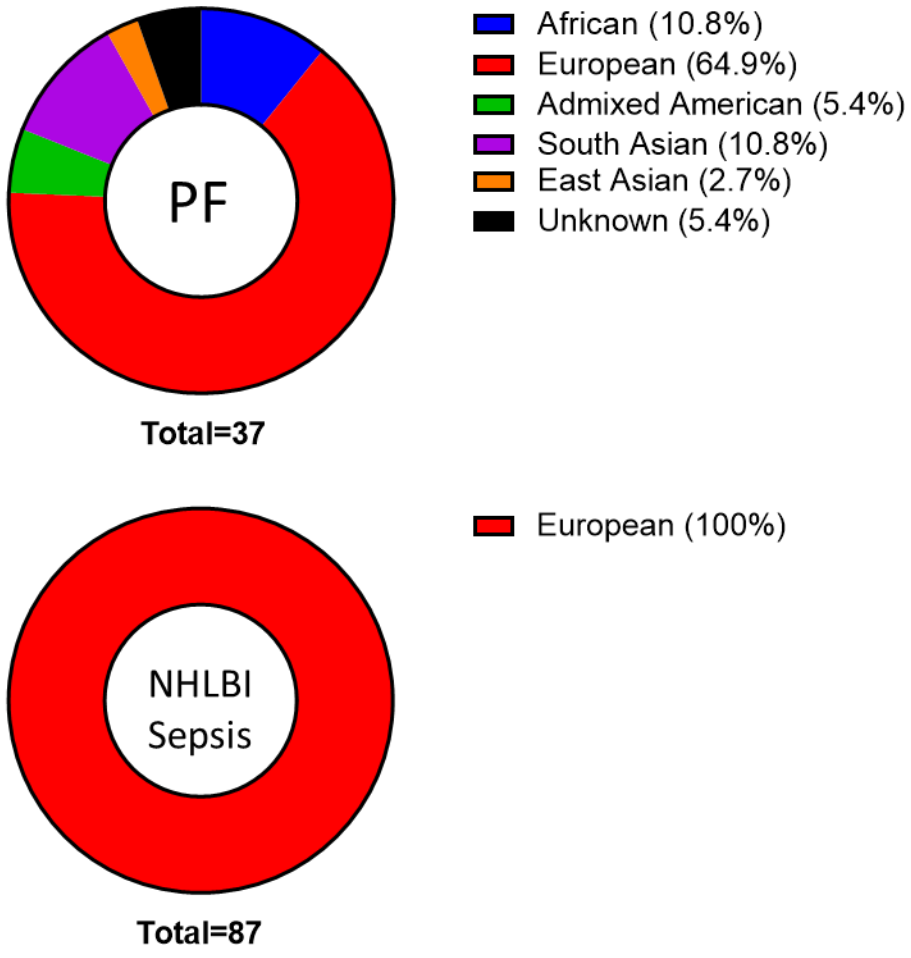

**Supplementary Figure 3: Burden of rare ΔP variants (MAF < 0.05) in the complement pathway with or without inclusion of non-European PF cases.**

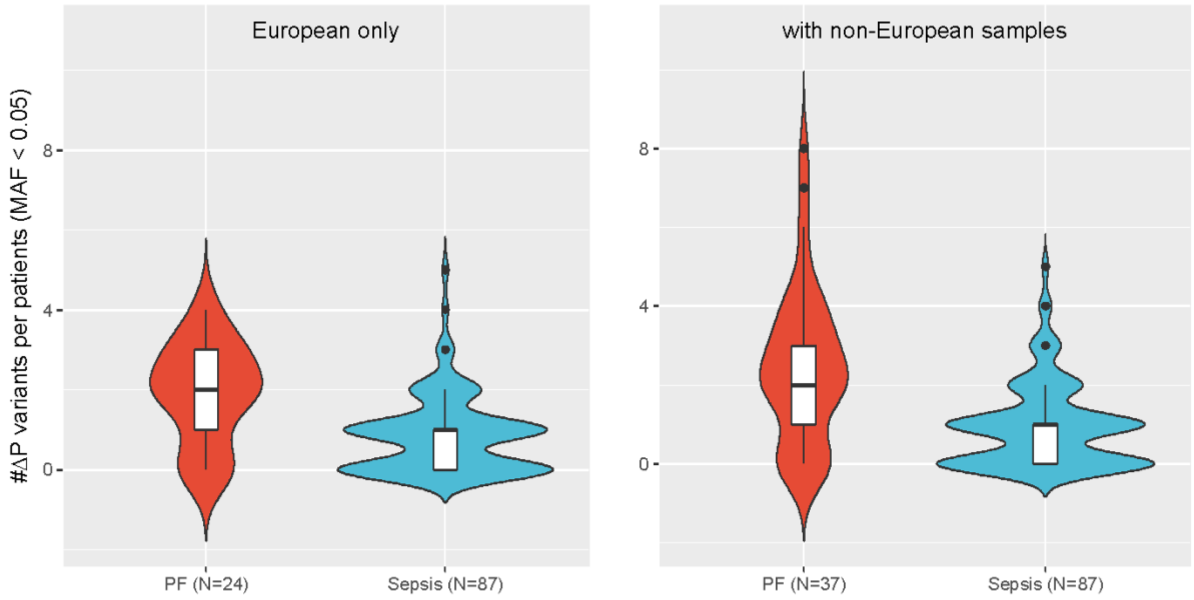

**Supplementary Figure 4: Complement receptor cloning strategy.** Each integrin subunit was co-expressed with a corresponding fluorescent reporter (GFP for the integrin beta chain and mCherry for the integrin alpha chain) as a single polypeptide chain. A P2A auto-cleavage site was placed between the fluorescent reporter and the integrin subunit such that, after cleavage at the P2A site, the fluorescent reporter was produced at a 1:1 molar ratio to the integrin subunit protein.

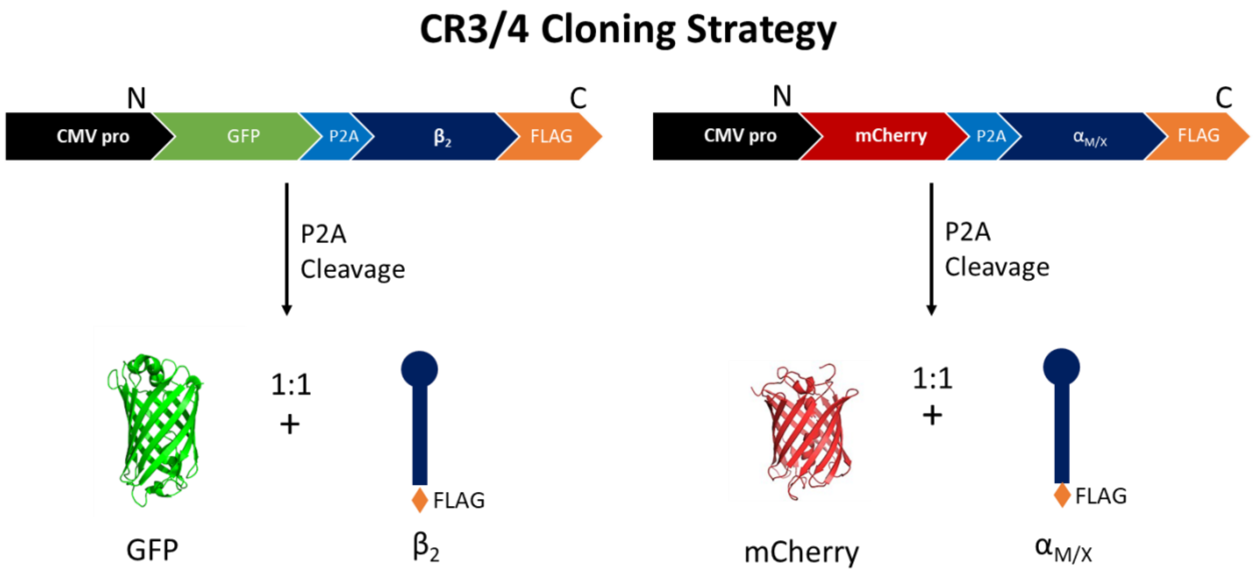

**Supplementary Figure 5: Expression of the CR3 and CR4 mutants.** HEK-293T cells were stably transfected with wild-type (WT) or mutant receptor subunits, along with their WT dimerization partners, and cell populations with equivalent GFP and mCherry double-positive fluorescence intensity were flow-sorted for each of the cell lines to ensure that each cell line expressed a similar amount of the transfected constructs. (**A**) Flow cytometry quantification of the relative expression of mCherry (linked to integrin alpha chain expression) and green fluorescent protein (GFP, linked to integrin beta chain expression), for each of the stable cell lines after sorting for similar mCherry and GFP fluorescence intensity. (**B**) Whole cell lysates from the indicated sorted cells were subjected to western blot analysis to quantify the total expression of each integrin subunit in each cell line. Untransfected (UT) HEK-293T cells were used as a negative control for integrin subunit expression. By sorting cells that had similar GFP and mCherry fluorescence, we ensured that the level of expression of each wild-type integrin protein and its corresponding PF-associated variants was similar across the different stable cell lines.

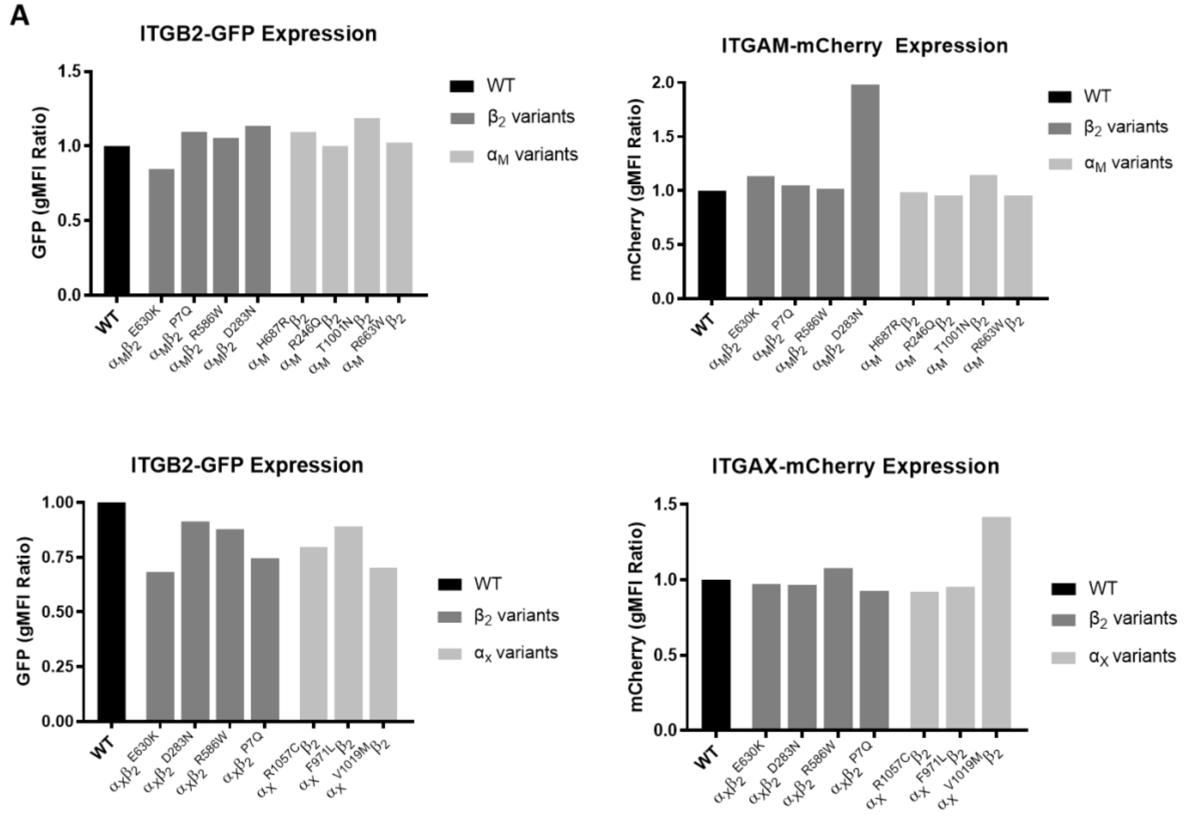

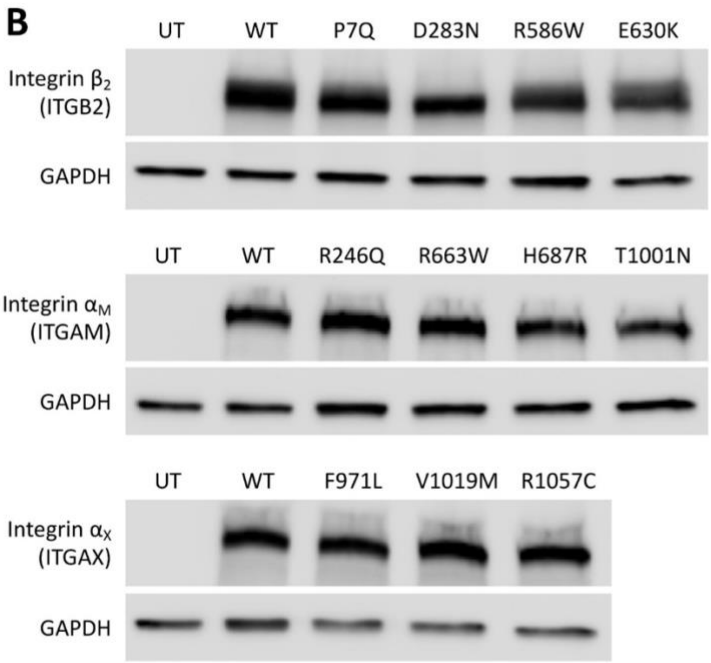
B
UT
WT
P7Q
D283N
R586W
E630K
Integrin β2 (ITGB2)
GAPDH
UT
WT
R246Q
R663W
H687R
T1001N
Integrin αM (ITGAM)
GAPDH
UT
WT
F971L
V1019M
R1057C
Integrin αX (ITGAX)
GAPDH

**Supplementary Table 1. Characteristics of the study cohorts.** Clinical and laboratory data at the time of presentation for patients in the Boston PF cohort (N=37) and the NHLBI ARDSnet iSPAAR control cohort (N=87).

| | **Boston PF Cohort (N=37)** | **NHLBI ARDSnet iSPAAR Cohort (N=87)** |
|---|---|---|
| Sex (% Male) | 59.5 | 70.1 |
| Mean age (±SD) | 44.5 ± 23.2 | 61.8 ± 17.2 |
| Race (% Caucasian) | 68.6 | 100% |
| Ventilated (%) | 80.0 | 81.6 |
| Mean platelet count (±SD) | 39.2 ± 35.2 | 203.4 ± 124.8 |
| Mean bilirubin (±SD) | 2.3 ± 2.4 | 1.3 ± 2.2 |
| Mean creatinine (±SD) | 3.6 ± 2.1 | 1.6 ± 1.1 |
| Mean lactate (±SD) | 9.0 ± 5.0 | UK |
| Mean protein C (% ±SD) | 22.8 ± 10.8 | UK |
| In-hospital mortality (%) | 42.4 | 21.8 |
| Mean SOFA score (±SD) | 12.8 ± 3.0 | 7.6 ± 2.8 |

UK: unknown

SD: standard deviation

SOFA: Sequential Organ Failure Assessment score

**Supplementary Table 2.** Infectious etiologies in the Boston PF Cohort (N=40).

| | |
|---|---|
| **Gram Negative (%)** | 18 (45) |
| *N. meningitidis* | 11 (27.5) |
| *C. canimorsus* | 4 (10.0) |
| *E. coli* | 2 (5.0) |
| *H. influenzae* | 1 (2.5) |
| **Gram Positive (%)** | 11 (27.5) |
| *S. pneumoniae* | 5 (12.5) |
| *S. aureus* | 2 (5.0) |
| Other strep spp. | 4 (10) |
| **Other/Unknown (%)** | 11 (27.5) |

**Supplementary Table 3. Test for distribution of doubletons and tripletons in the European PF cases (N=24) and unselected sepsis controls (N=87).** Doubletons refer to variants with allele counts of 2 and tripletons refer to variants with allele counts of 3 in the study cohort.

| Type | Frequency | PF only (N=24) | Sepsis only (N=87) | Shared | Binomial test p-value (one-tailed) |
|---|---|---|---|---|---|
| Synonymous | Doubletons | 33 | 148 | 105 | 0.857491446 |
| | Tripletons | 6 | 67 | 75 | 0.516643659 |
| Missense | Doubletons | 56 | 220 | 156 | 0.847182549 |
| | Tripletons | 13 | 115 | 137 | 0.633240968 |

**Supplementary Table 4. Pathway gene sets used for statistical analyses.**

| Pathway | Genes |
|---|---|
| Complement (N=27) | C1QA, C1QB, C1QC, C1R, C1S, C2, C3, C3AR1, C4A, C4B, C5, C5AR1, C6, C7, C8A, C8B, C8G, C9, CFB, CFD, CFH, CFI, CFP, CR2, ITGAM, ITGAX, ITGB2 |
| Coagulation (N=45) | A2M, BDKRB1, BDKRB2, F10, F11, F12, F13A1, F13B, F2, F2R, F2RL2, F2RL3, F3, F5, F7, F8, F9, FGA, FGB, FGG, KLKB1, KNG1, MASP1, MASP2, MBL2, PLAT, PLAU, PLAUR, PLG, PROC, PROCR, PROS1, SERPINA1, SERPINA5, SERPINB2, SERPINC1, SERPIND1, SERPINE1, SERPINF2, SERPING1, TFPI, THBD, VSIG4, VTN, VWF |
| Glycolysis (N=68) | HK3, HK1, HK2, HKDC1, GCK, GPI, PFKM, PFKP, PFKL, FBP1, FBP2, ALDOC, ALDOA, ALDOB, TPI1, GAPDH, GAPDHS, PGK2, PGK1, PGAM1, PGAM2, PGAM4, ENO3, ENO2, ENO1, ENO4, PKM, PKLR, PDHA2, PDHA1, PDHB, DLAT, DLD, LDHAL6A, LDHAL6B, LDHA, LDHB, LDHC, ADH1A, ADH1B, ADH1C, ADH7, ADH4, ADH5, ADH6, AKR1A1, ALDH2, ALDH3A2, ALDH1B1, ALDH7A1, ALDH9A1, ALDH3B1, ALDH3B2, ALDH1A3, ALDH3A1, ACSS1, ACSS2, GALM, PGM1, PGM2, G6PC, G6PC2, G6PC3, ADPGK, BPGM, MINPP1, PCK1, PCK2 |

**Supplementary Table 5. All rare non-synonymous variants identified in the humoral complement system sub-pathway.** Known or predicted functional consequences of each variant and the number of PF and unselected sepsis patients with the variant are indicated. Variants with global MAF < 0.05 in gnomAD database were considered to be rare. SDV: Splice Donor Variant; SAV: Splice Acceptor Variant; LoF: Loss-of-function; INS: Insertion.

| Humoral Variants | Chromosome: Position | Nucleotide change | # in PF | # in sepsis | Known/Predicted effect |
|---|---|---|---|---|---|
| C1R INS | 12:7188256 | A>AACGATAC | 2 | 0 | ΔP – high-precision prediction LoF |
| C1R G359E | 12:7241090 | C>T | 1 | 0 | predicted neutral |
| C1R A140V | 12:7242657 | G>A | 0 | 1 | ΔP - predicted consequential (both) |
| C1R M112I | 12:7242740 | C>G | 0 | 1 | ΔP - predicted consequential (both) |
| C1R G93C | 12:7242799 | C>A | 1 | 0 | ΔP - predicted consequential (both) |
| C1R E11X | 12:7244382 | C>A | 0 | 1 | ΔP – high-precision prediction LoF |
| C1S SRV | 12:7169990 | T>G | 1 | 0 | ΔP – splice region variant |
| C1S SRV | 12:7169992 | T>G | 1 | 0 | ΔP – splice region variant |
| C1S SRV | 12:7169993 | T>G | 1 | 0 | ΔP – splice region variant |
| C1S A307V | 12:7173870 | C>T | 1 | 0 | predicted neutral |
| C1S D315N | 12:7173893 | G>A | 0 | 1 | ΔP - predicted consequential (both) |
| C1S R383H | 12:7175028 | G>A | 1 | 0 | predicted neutral |
| C2 N27K | 6:31895766 | C>G | 0 | 1 | ΔP - predicted consequential (PolyPhen2) |
| C2 P73S | 6:31895902 | C>T | 1 | 1 | predicted neutral |
| C2 c.841_849+19delGTGGACAGGGTCAGGAATCAGGAGTCTG | 6:31902065 | ATGGTGGACAGGGTCAGGAATCAGGAGTC>A | 0 | 1 | ΔP – SDV: high-precision prediction LoF |
| C2 E318D | 6:31903804 | G>C | 4 | 3 | ΔP - predicted consequential (PolyPhen2) and consequential in prior literature[1,2] |
| C2 V641A | 6:31912523 | T>C | 1 | 1 | predicted neutral |
| C3 L1549M | 19:6678452 | G>T | 1 | 0 | ΔP - predicted consequential (SIFT) |
| C3 D1440A | 19:6681983 | T>G | 0 | 1 | predicted neutral |
| C3 c.3970-8C>T | 19:6684853 | G>A | 1 | 0 | ΔP – splice region variant |
| C3 R735W | 19:6707129 | G>A | 1 | 1 | ΔP - predicted consequential (both) and consequential in prior literature[3] |

| | | | | | |
|---|---|---|---|---|---|
| C3 INS | 19:6711212 | G>GACTCCT | 1 | 0 | ΔP – splice region variant |
| C3 K155Q | 19:6718146 | T>G | 0 | 1 | predicted neutral |
| C3AR1 L333P | 12:8211784 | A>G | 2 | 3 | ΔP - predicted consequential (SIFT) |
| C3AR1 V136A | 12:8212375 | A>G | 1 | 0 | ΔP - predicted consequential (SIFT) |
| C3AR1 c.-8G>A | 12:8212789 | C>T | 0 | 1 | ΔP – splice region variant |
| C4A P1120T | 6:31963859 | C>A | 0 | 1 | ΔP - predicted consequential (PolyPhen2) |
| C4B c.3505-8T>C | 6:31996936 | T>C | 1 | 2 | ΔP – splice region variant |
| C5 INS | 9:123716017 | A>AGTAT | 3 | 0 | ΔP – high-precision prediction LoF |
| C5 SRV | 9:123739189 | C>CTTGAAAG | 1 | 0 | ΔP – splice region variant |
| C5 SAV | 9:123777540 | C>G | 1 | 0 | ΔP – SAV: high-precision prediction LoF |
| C5 SRV | 9:123777543 | G>A | 1 | 0 | ΔP – splice region variant |
| C5 SRV | 9:123777546 | T>G | 1 | 0 | ΔP – splice region variant |
| C5 L571V | 9:123779926 | G>C | 0 | 1 | ΔP - predicted consequential (both) |
| C5 INS | 9:123785771 | G>GGATTCCA | 1 | 0 | ΔP – high-precision prediction LoF |
| C5AR1 R320W | 19:47823992 | C>T | 0 | 1 | ΔP - predicted consequential (SIFT) |
| C5AR1 T342A | 19:47824058 | A>G | 0 | 1 | ΔP - predicted consequential (SIFT) |
| C6 A934T | 5:41142932 | C>T | 1 | 0 | predicted neutral |
| C6 G840S | 5:41149448 | C>T | 1 | 0 | predicted neutral |
| C6 c.2381+2T>C | 5:41150035 | A>G | 1 | 0 | ΔP – SDV: high-precision prediction LoF and consequential in prior literature[4] |
| C6 c.2101+3G>T | 5:41155071 | C>A | 0 | 1 | ΔP – splice region variant |
| C6 D696G | 5:41155088 | T>C | 1 | 3 | predicted neutral |
| C6 D667H | 5:41155176 | C>G | 0 | 1 | predicted neutral |
| C6 M616I | 5:41159192 | C>T | 1 | 0 | ΔP - predicted consequential (SIFT) |
| C6 Q567H | 5:41159339 | C>G | 1 | 0 | predicted neutral |
| C6 D519N | 5:41160373 | C>T | 2 | 0 | ΔP - predicted consequential (both) |
| C6 S442P | 5:41161929 | A>G | 1 | 0 | ΔP - predicted consequential (both) |
| C6 I217T | 5:41186248 | A>G | 0 | 1 | predicted neutral |
| C6 T181I | 5:41195939 | G>A | 0 | 1 | predicted neutral |
| C6 N74H | 5:41201740 | T>G | 0 | 1 | ΔP - predicted consequential (both) |
| C6 R4L | 5:41203322 | C>A | 1 | 0 | predicted neutral |
| C7 C128R | 5:40936541 | T>C | 0 | 2 | ΔP - predicted consequential (both) |
| C7 K420Q | 5:40955653 | A>C | 1 | 0 | ΔP - predicted consequential (both) |
| C7 R521S | 5:40959622 | C>A | 1 | 0 | ΔP - predicted consequential (both) and consequential in prior literature[5] |

| | | | | | |
|---|---|---|---|---|---|
| C7 R618W | 5:40964945 | C>T | 0 | 1 | ΔP - predicted consequential (both) |
| C7 A670T | 5:40972630 | G>A | 0 | 1 | predicted neutral |
| C8A A36E | 1:57333311 | C>A | 3 | 3 | ΔP - predicted consequential (both) |
| C8A E320D | 1:57351704 | G>T | 0 | 1 | predicted neutral |
| C8A R444H | 1:57373737 | G>A | 1 | 0 | ΔP - predicted consequential (both) |
| C8A D458N | 1:57373778 | G>A | 1 | 0 | ΔP - predicted consequential (both) |
| C8A P475R | 1:57378119 | C>G | 1 | 0 | predicted neutral |
| C8A P575S | 1:57383357 | C>T | 2 | 2 | predicted neutral |
| C8B T542I | 1:57395228 | G>A | 0 | 2 | predicted neutral |
| C8B R428X | 1:57406638 | G>A | 0 | 1 | ΔP – high-precision prediction LoF |
| C8B D382Y | 1:57409459 | C>A | 0 | 2 | ΔP - predicted consequential (both) |
| C8B H277D | 1:57415263 | G>C | 0 | 1 | ΔP - predicted consequential (SIFT) |
| C8B P261L | 1:57415310 | G>A | 4 | 8 | predicted neutral |
| C8B R242H | 1:57415367 | C>T | 0 | 1 | predicted neutral |
| C8B T220K | 1:57417728 | G>T | 1 | 0 | ΔP - predicted consequential (SIFT) |
| C8B G198A | 1:57417794 | C>G | 0 | 1 | ΔP - predicted consequential (both) |
| C8G p.Ser34ProfsTer12 | 9:139839860 | G>GCATCCCC | 0 | 1 | ΔP – high-precision prediction LoF |
| C8G c.276-3C>T | 9:139840378 | C>T | 0 | 1 | ΔP – splice region variant |
| C9 F338L | 5:39311336 | A>T | 0 | 1 | ΔP - predicted consequential (both) |
| C9 T279S | 5:39315911 | G>C | 1 | 0 | predicted neutral |
| C9 P167S | 5:39331894 | G>A | 0 | 2 | ΔP - predicted consequential (both) |
| C9 M45L | 5:39342243 | T>A | 0 | 1 | predicted neutral |
| CFB L9H | 6:31914024 | T>A | 4 | 8 | ΔP - predicted consequential (both) and consequential in prior literature[2] |
| CFB G252S | 6:31915614 | G>A | 3 | 8 | predicted neutral |
| CFB T882P | 6:31916708 | A>C | 0 | 3 | ΔP - predicted consequential (both) |
| CFB c.1408+7A>C | 6:31917341 | A>C | 2 | 0 | ΔP – splice region variant |
| CFB K1067E | 6:31918464 | A>G | 1 | 0 | ΔP - predicted consequential (both) |
| CFB E1068A | 6:31918468 | A>C | 1 | 0 | predicted neutral |
| CFD E69K | 19:860766 | G>A | 1 | 2 | predicted neutral |
| CFD c.212+2T>G | 19:860775 | T>G | 2 | 0 | ΔP – SDV: high-precision prediction LoF |
| CFH c.245-7G>A | 1:196642980 | G>A | 2 | 0 | ΔP – splice region variant |
| CFH I551T | 1:196684855 | T>C | 1 | 0 | ΔP - predicted consequential (both) |
| CFH c.2236+8T>A | 1:196696078 | T>A | 1 | 1 | ΔP – splice region variant |
| CFH S890I | 1:196706677 | G>T | 3 | 0 | predicted neutral |

| | | | | | |
|---|---|---|---|---|---|
| CFH Q950H | 1:196709816 | G>T | 0 | 1 | ΔP - predicted consequential (SIFT) |
| CFH T956M | 1:196709833 | C>T | 0 | 1 | predicted neutral |
| CFH V1007L | 1:196711067 | G>T | 4 | 0 | predicted neutral |
| CFH T1038R | 1:196711161 | C>G | 0 | 1 | ΔP - predicted consequential (SIFT) |
| CFH N1050Y | 1:196712596 | A>T | 0 | 2 | predicted neutral |
| CFH I1059T | 1:196712624 | T>C | 1 | 0 | ΔP - predicted consequential (SIFT) |
| CFH Q1143E | 1:196715063 | C>G | 1 | 0 | predicted neutral |
| CFH R1203W | 1:196716354 | C>T | 0 | 1 | predicted neutral |
| CFI c.1534+5G>T | 4:110663642 | C>A | 2 | 1 | ΔP – splice region variant |
| CFI K441R | 4:110667485 | T>C | 1 | 2 | predicted neutral |
| CFI I416L | 4:110667561 | T>G | 0 | 1 | predicted neutral |
| CFI R406H | 4:110667590 | C>T | 1 | 1 | predicted neutral |
| CFI A240G | 4:110681732 | G>C | 1 | 0 | ΔP - predicted consequential (both) |
| CFI R187X | 4:110682772 | G>A | 1 | 0 | ΔP – high-precision prediction LoF |
| CFI Q161H | 4:110682848 | T>A | 1 | 0 | ΔP - predicted consequential (PolyPhen2) |
| CFI c.482+8C>T | 4:110685685 | G>A | 0 | 1 | ΔP – splice region variant |
| CFI c.482+6C>A | 4:110685687 | G>T | 1 | 0 | ΔP – splice region variant |
| CFI c.482+6C>T | 4:110685687 | G>A | 2 | 0 | ΔP – splice region variant |
| CFP T400A | X:47485503 | T>C | 0 | 1 | predicted neutral |

**Supplementary Table 6. All rare non-synonymous variants identified in the opsonophagocytic complement system sub-pathway.** Known or predicted functional consequences of each variant and the number of PF and unselected sepsis patients with the variant are indicated. Variants with a Gnomad global MAF of < 0.05 were considered to be rare. SDV: Splice Donor Variant; SAV: Splice Acceptor Variant; LoF: Loss-of-function; INS: Insertion.

| Opsonophagocytic Variants | Chromosome: Position | Nucleotide change | # in PF | # in sepsis | Known/Predicted effect |
|---|---|---|---|---|---|
| CR2 V130A | 1:207640201 | T>C | 1 | 0 | ΔP - predicted consequential (SIFT) |
| CR2 N223S | 1:207642178 | A>G | 0 | 1 | ΔP - predicted consequential (both) |
| CR2 P307L | 1:207643142 | C>T | 0 | 1 | ΔP - predicted consequential (both) |
| CR2 T334I | 1:207643223 | C>T | 0 | 1 | predicted neutral |
| CR2 P404A | 1:207643432 | C>G | 0 | 1 | ΔP - predicted consequential (both) |
| CR2 R514W | 1:207646163 | C>T | 2 | 2 | ΔP - predicted consequential (SIFT) |
| CR2 S541I | 1:207646168 | G>T | 2 | 0 | ΔP - predicted consequential (SIFT) |
| CR2 G559E | 1:207646222 | G>A | 0 | 1 | ΔP - predicted consequential (both) |
| CR2 c.1978+7A>T | 1:207646531 | A>T | 0 | 1 | ΔP - splice region variant |
| CR2 H669R | 1:207646917 | A>G | 0 | 1 | ΔP - predicted consequential (both) |
| CR2 V812L | 1:207648456 | G>T | 0 | 1 | ΔP - predicted consequential (SIFT) |
| CR2 Q902L | 1:207649744 | A>T | 0 | 1 | predicted neutral |
| CR2 Q1011H | 1:207651360 | G>C | 1 | 0 | ΔP - predicted consequential (both) |
| CR2 c.3189-8_3189-7delCT | 1:207658799 | TTC>T | 1 | 0 | ΔP - splice region variant |
| ITGAM R246Q | 16:31284718 | G>A | 1 | 0 | ΔP - predicted consequential (both) |
| ITGAM K247E | 16:31284720 | A>G | 0 | 1 | predicted neutral |
| ITGAM R415W | 16:31289317 | C>T | 0 | 1 | ΔP - predicted consequential (SIFT) |
| ITGAM c.1497+1dupG | 16:31308970 | A>AG | 0 | 1 | ΔP – high-precision prediction LoF |
| ITGAM R663W | 16:31332930 | C>T | 1 | 0 | ΔP - predicted consequential (SIFT) |
| ITGAM R663Q | 16:31332931 | G>A | 0 | 1 | predicted neutral |
| ITGAM H687R | 16:31335774 | A>G | 1 | 0 | predicted neutral |
| ITGAM Ins | 16:31336642 | A>AGAAG | 3 | 0 | ΔP – high-precision prediction LoF |
| ITGAM Q955H | 16:31340618 | A>C | 0 | 1 | ΔP - predicted consequential (PolyPhen2) |
| ITGAM T1001N | 16:31341424 | C>A | 1 | 0 | predicted neutral |

| | | | | | |
|---|---|---|---|---|---|
| ITGAX G130R | 16:31368643 | G>C | 0 | 1 | ΔP - predicted consequential (both) |
| ITGAX I286T | 16:31371780 | T>C | 0 | 1 | ΔP - predicted consequential (both) |
| ITGAX R750W | 16:31383786 | C>T | 0 | 2 | ΔP - predicted consequential (SIFT) |
| ITGAX F971L | 16:31391120 | T>C | 1 | 0 | ΔP - predicted consequential (SIFT) |
| ITGAX V1019M | 16:31391381 | G>A | 1 | 1 | predicted neutral |
| ITGAX R1057C | 16:31391695 | C>T | 1 | 0 | ΔP - predicted consequential (SIFT) |
| ITGAX c.3388-5C>T | 16:31393119 | C>T | 0 | 1 | ΔP - splice region variant |
| ITGB2 P7Q | 21:46330678 | G>T | 1 | 0 | predicted neutral |
| ITGB2 R120W | 21:46323421 | G>A | 0 | 1 | ΔP - predicted consequential (both) |
| ITGB2 P201L | 21:46321546 | G>A | 0 | 1 | ΔP - predicted consequential (both) |
| ITGB2 D283N | 21:46320285 | C>T | 1 | 0 | ΔP - predicted consequential (both) |
| ITGB2 R456C | 21:46311770 | G>A | 0 | 1 | ΔP - predicted consequential (both) |
| ITGB2 R586W | 21:46309312 | G>A | 1 | 0 | ΔP - predicted consequential (PolyPhen2) and consequential in prior literature[6] |
| ITGB2 E630K | 21:46308800 | C>T | 2 | 0 | ΔP - predicted consequential (both) |
| ITGB2 c.499+7C>T | 21:46323273 | G>A | 1 | 2 | ΔP - splice region variant |
| ITGB2 c.1412+8G>A | 21:46311716 | C>T | 1 | 0 | ΔP - splice region variant |

**Supplementary Table 7. Rare variant burden of complement system variants.** Variant burden was assessed by one-sided Fisher's exact test on the European PF cases (N=24) and the unselected sepsis controls (N=87).

| Type | Cases with | Controls with | Cases without | Controls without | Fisher's one-tailed p-value |
|---|---|---|---|---|---|
| ΔP variants | 18 | 46 | 6 | 41 | **0.0418** |
| Neutral Missense | 16 | 66 | 8 | 21 | 0.2551 |
| Synonymous | 15 | 40 | 9 | 47 | 0.1144 |

**Supplementary Table 8. Gene-based collapsing test on complement system genes in quality-filtered European PF cases and sepsis controls.** Genes with no qualifying variants are not shown.

| Gene | CMC Fisher p-value | CMC Fisher p-value | Kbac p-value | Kbac p-value | SKAT perm p-value | SKAT perm p-value (Bonferonni corrected) | VTPrice perm p-value | VTPrice perm p-value (Bonferroni corrected) |
|---|---|---|---|---|---|---|---|---|
| C1R | 0.0825 | 1.0000 | 0.1104 | 1.0000 | 0.0579 | 1.0000 | 0.0671 | 1.0000 |
| C1S | 0.5978 | 1.0000 | 1.0000 | 1.0000 | 0.6001 | 1.0000 | 0.6145 | 1.0000 |
| C2 | 0.0827 | 1.0000 | 0.0727 | 1.0000 | 0.0271 | 0.7304 | 0.1013 | 1.0000 |
| C3 | 0.3252 | 1.0000 | 1.0000 | 1.0000 | 0.3279 | 1.0000 | 0.3697 | 1.0000 |
| C3AR1 | 0.3070 | 1.0000 | 1.0000 | 1.0000 | 0.3130 | 1.0000 | 0.6240 | 1.0000 |
| C4A | 0.5978 | 1.0000 | 1.0000 | 1.0000 | 0.5917 | 1.0000 | 0.6170 | 1.0000 |
| C5 | 0.3252 | 1.0000 | 0.3903 | 1.0000 | 0.1397 | 1.0000 | 0.2177 | 1.0000 |
| C5AR1 | 0.4535 | 1.0000 | 1.0000 | 1.0000 | 0.7525 | 1.0000 | 0.6885 | 1.0000 |
| C6 | 0.0547 | 1.0000 | 0.2026 | 1.0000 | 0.0567 | 1.0000 | 0.5003 | 1.0000 |
| C7 | 0.8672 | 1.0000 | 1.0000 | 1.0000 | 0.2536 | 1.0000 | 0.1973 | 1.0000 |
| C8A | 0.0825 | 1.0000 | 0.1142 | 1.0000 | 0.1514 | 1.0000 | 0.1170 | 1.0000 |
| C8B | 0.6588 | 1.0000 | 0.7896 | 1.0000 | 0.9158 | 1.0000 | 0.6547 | 1.0000 |
| C8G | 0.5978 | 1.0000 | 1.0000 | 1.0000 | 0.5970 | 1.0000 | 0.0725 | 1.0000 |
| C9 | 0.3564 | 1.0000 | 1.0000 | 1.0000 | 0.6353 | 1.0000 | 0.7869 | 1.0000 |
| CFB | 0.2108 | 1.0000 | 0.0669 | 1.0000 | 0.0656 | 1.0000 | 0.0925 | 1.0000 |
| CFI | 0.0066 | 0.1778 | 0.2312 | 1.0000 | 0.0255 | 0.6884 | 0.0033 | 0.0891 |
| CR2 | 0.6315 | 1.0000 | 1.0000 | 1.0000 | 0.4208 | 1.0000 | 0.5120 | 1.0000 |
| ITGAM | 0.8672 | 1.0000 | 1.0000 | 1.0000 | 0.3482 | 1.0000 | 0.3440 | 1.0000 |
| ITGAX | 0.2847 | 1.0000 | 1.0000 | 1.0000 | 0.7517 | 1.0000 | 0.6329 | 1.0000 |
| ITGB2 | 0.0183 | 0.4954 | 0.0200 | 0.5399 | 0.0085 | 0.2295 | 0.0236 | 0.6359 |

**Methods:**

**Study samples**

The Boston PF Cohort is a multi-institutional dataset comprised of PF patients who presented to three large academic medical centers in Boston or whose cases were referred to one of these institutions, between 1995 and 2019. Cases meeting PF criteria (see below) that had specimens suitable for next generation sequencing (NGS) were selected (N=40). Given the rarity of PF, we obtained approximately a third of our samples by extracting genomic DNA from archived formalin-fixed, paraffin-embedded (FFPE) surgical pathology specimens. The remaining samples were comprised of genomic DNA extracted from whole blood or cheek swab specimens from living subjects. Archived FFPE tissue was obtained via an automated pipeline that leveraged deep-mining of the electronic medical record systems at the three Boston sites. Using natural language processing, we searched for the term “purpura fulminans” in patient charts dating back to the 1990s. Targeted manual chart review was then used on candidate medical records to identify those meeting the case definition of PF. Cases meeting the definition of PF with available FFPE tissue from autopsies or premortem surgical pathology specimens were included in the cohort for germline sequencing. A group of unselected patients with sepsis from the NHLBI ARDSnet iSPAAR consortium contained in the NIH Database of Genotypes and Phenotypes (dbGaP) was used as a control cohort (dbGaP study accession phs000631.v1.p1, N=87). Exome sequences for these 87 patients were downloaded directly from DBGaP. This study was approved by the institutional review boards at all participating sites.

**Case definition for Purpura Fulminans**

Each case was adjudicated by a panel of three experts prior to inclusion in the study. PF was defined as the rapid onset of systemic inflammatory response syndrome, evidence of consumptive coagulopathy, and skin findings suggestive of purpura. One or more of the following was used to define consumptive coagulopathy:

1. Activated partial thromboplastin time (aPTT) ≥ 45 seconds

2. International Normalized Ratio ≥ 1.5

3. Platelets ≤ 100,000 per μL

4. D-dimer ≥ 3000 ng/mL

Patients with purpura fulminans were more likely to also have the following:

1. Elevated lactate

2. Low or inappropriately normal fibrinogen

3. Elevated C-reactive protein (CRP) with inappropriately low erythrocyte sedimentation rate (ESR)

4. Dog owner (infection with *Capnocytophaga canimorsus*)

5. Meningeal signs and/or history suggestive of *Neisseria meningitidis* infection

6. Immunocompromised state, including HIV, alcoholism, or asplenia

7. Low levels of endogenous anticoagulant proteins (*e.g.* proteins C/S and antithrombin)

**Whole exome sequencing**

For the PF patient samples, germline genomic DNA was extracted from either whole blood or formalin-fixed, paraffin-embedded tissue samples. Sequencing and variant calling were performed at the Yale Center for Genomic Analysis (New Haven, CT). Paired-end sequencing was carried out on the Illumina HiSeq 4000 to a minimum coverage depth of 20X.

*Library construction*

Library construction was performed as previously described[7], with the following modifications: initial genomic DNA input into shearing was reduced from 3 µg to 10-100 ng in 50 µL of solution. For adapter ligation, paired-end adapters (Illumina; San Diego, CA, USA) were replaced with palindromic forked adapters (Integrated DNA Technologies; Coralville, IA, USA) with unique dual-indexed molecular barcode sequences to facilitate downstream pooling. With the exception of the palindromic forked adapters, all of the reagents used for end-repair, A-base addition, adapter ligation, and library enrichment

PCR were purchased from KAPA Biosystems (Wilmington, MA, USA). In addition, during the post-enrichment SPRI cleanup, elution volume was reduced to 30 µL to maximize library concentration and a vortexing step was added to maximize the amount of template eluted.

*In-solution hybrid selection*

After library construction, hybridization and capture were performed using the relevant components of the Nextera Rapid Capture Exome Kit (Illumina) following the manufacturer's suggested protocol, with the following exceptions: first, all libraries within a library construction plate were pooled prior to hybridization. Second, the Midi plate from the Nextera Rapid Capture Exome Kit was replaced with a skirted PCR plate to facilitate automation. All hybridization and capture steps were automated on the Bravo liquid handling system (Agilent Technologies; Santa Clara, CA, USA).

*Preparation of libraries for cluster amplification and sequencing*

After post-capture enrichment, library pools were quantified by automated qPCR assay on the Agilent Bravo using a kit purchased from KAPA Biosystems with probes specific to the ends of the adapters. Based on qPCR quantification, libraries were normalized to 2 nM, then denatured using 0.1 N NaOH on the STARlet liquid handler (Hamilton Microlab; Reno, NV, USA). After denaturation, libraries were diluted to 20 pM using hybridization buffer (Illumina).

*Cluster amplification and sequencing*

Cluster amplification of denatured templates was performed according to the manufacturer's protocol (Illumina) using HiSeq 4000 cluster chemistry and HiSeq 4000 flowcells. Flowcells were sequenced by v1 Sequencing-by-Synthesis chemistry. The flowcells were then analyzed using RTA v.1.18.64 or later. Each pool of whole exome libraries was run on paired 76 base pair runs reading the dual-indexed sequences to identify molecular indices and was sequenced across the number of lanes needed to meet coverage (mean target coverage depth >20X).

*Data processing, filtering and variant calling*

The sequencing reads were analyzed by an exome analysis pipeline that follows GATK 3.5 best practices workflow for alignment and variant calling. Reads were first aligned to the hg19 human reference with decoy sequences (hs37d5) using BWA MEM v0.7.15. PCR duplicates were marked using Picard's MarkDuplicates v2.8.2. Then, GATK 3.5 software was used to perform indel realignment, base quality score recalibration, and generation of GVCF files using HaplotypeCaller. The target regions used for variant calling were the union of the Integrated DNA Technologies xGen exome kit target regions padded by 40 base pairs on either side, plus all the RefGene coding regions padded by 15 base pairs on either side. Once the gVCF file was generated, the variants were jointly called across the PF and unselected sepsis cohort. GATK's hard filtering commands were used to filter the variants. We initially retained variants that had a missing genotype rate of <= 10% and minimum read depth >= 10x. Among those, we kept SNPs that were "PASS" variants with genotype call quality ≥ 90%, and indels that were "VQSRTrancheINDEL99.00to99.90" with allele balance ≥ 25%. Missing genotypes were not imputed.

**Data Analysis**

*Sample quality control*

Principal component analysis (PCA) was performed using common, known SNPs (MAF >= 1%) from the PF cases and sepsis controls (**Supplementary Figure 2**). Among the cases, 67.5% were European (N=27), 10% were African, 10% were South Asian, 5% were admixed American, 2.5% were East Asian, and the rest were of unknown ethnicity. To ensure that our results were not driven or confounded by population structure, we performed burden analysis on European-only cases and controls. Additionally, we computed per sample Ti/Tv ratios and removed outliers that were more than three standard deviations from the exome-wide average Ti/Tv ratio for all the samples (**Supplementary Figure 1**). Three European samples (BGM0330, BGM0336, and BGM0338) were found to be outliers not only in terms of Ti/Tv ratio, but also in terms of Het/Hom ratio and total SNVs per haploid genome. They also showed an unusually large number of transversions in novel SNPs compared to transition suggesting the presence of putative false positive missense mutations. After removing the anomalous samples, 24 European PF cases were retained for further analysis. All of the unselected control sepsis samples passed the quality control steps and were included in the analysis.

*Variant quality control*

To control for putative false positive variants, we performed a conservative analysis considering only bi-allelic SNPs and indels within the canonical transcripts of protein-coding genes for this study. Additionally, conforming to the GATK best practices, we included only those bi-allelic SNPs that have quality by depth (QD) > 2.0, Phred-scaled probability of strand bias (FS) < 60, strand odds ratio (SOR) <= 3.0, z-approximation from the Rank Sum Test for mapping qualities (MQRankSum) > -12.5, and z-approximation from the Rank Sum Test for site position within reads (ReadPosRankSum) > -8.0. As for indels, we included the ones having QD > 2.0, FS < 200, SOR < 10.0, and ReadPosRankSum > -20.0. Additionally, we excluded any variant (SNP or indel) having an inbreeding coefficient <= -0.8.

We computed doubletons (bi-allelic SNPs for which the alternative allele is observed only twice in the population) and tripletons (bi-allelic SNPs for which the alternative allele is observed only thrice in the population) in all the retained samples to ensure that their distribution follows the binomial expectation. This test enables us to examine whether the distribution of ultra-rare alleles in the case-control cohorts is driven by the underlying structure of the data. We detected no differences when evaluating the distribution of doubletons and tripletons that are a) shared between PF and sepsis individuals, b) specific to either PF or sepsis individuals, a finding that further supports the absence of substructure in the dataset (**Supplementary Table 3**).

*Gene sets*

A preliminary manual review of candidate variants was performed using Forome AnFiSA. We then performed pathway-based rare variant analysis on two pathways of interest, the complement system and the coagulation system, both of which are highly relevant to PF pathology. We also used the glycolysis pathway gene set as a control. Complete gene sets are listed in **Supplementary Table 4**. Constituents of the complement system gene set were chosen a priori based on review of the scientific literature with a focus on complement proteins important for clearance of encapsulated organisms and/or relevant to regulation of inflammation. The coagulation gene set was selected to contain pro-coagulant proteins as well as those that are required for negative regulation of coagulation. For the glycolysis pathway, the gene set listed in the Broad Institute's MSigDB database was used (https://www.gsea-msigdb.org/gsea/msigdb/).

**Statistical analysis of rare variant burden in PF**

We developed a novel approach to assessing the relationship between the increasing burden of qualifying rare variants in our pathway of interest with disease status by using our pathway-based rare variant trend test (RVTT). This analysis was motivated by the observation that very few individuals lack a single rare coding variant in a large pathway limiting the power of traditional burden tests focusing on presence vs. absence of rare variants.

We treated the burden of variants as an ordinal variable and disease status as a binary variable. Suppose qualifying rare variants occur in the pathway of interest 1, 2, …, I times, and the disease status is encoded as 1 (= PF) or 0 (= Sepsis). At any MAF cutoff, we can summarize the counts of qualifying rare variants in cases and controls using a 2xI contingency table with the ordered columns indicating the number of occurrences of qualifying rare variants i and the rows indicating the binary disease status.

| Disease Status | # Occurrences of qualifying rare variants in pathway | | | |
|---|---|---|---|---|
| | 1 | 2 | i | I |
| 1 (= PF) | $n_{11}$ | $n_{12}$ | $n_{1i}$ | $n_{1I}$ |
| 0 (= Sepsis) | $n_{01}$ | $n_{02}$ | $n_{0i}$ | $n_{0I}$ |

Here, the Null hypothesis (H0) is that there is no linear trend in binomial proportions of disease status across increasing numbers of qualifying variants in the pathway of interest; and the Alternative hypothesis (H1) is that there is a linear trend in binomial proportions of disease status across increasing numbers of qualifying variants in the pathway of interest. Let $p_{1|i}$ denote the probability of PF and $x_{1|i}$ denote the proportion of observed PF samples with i qualifying rare variants in the pathway of interest, where $i = 1, 2, \ldots, I$.

For the linear probability model,

$$p_{1|i} = \alpha + \beta * i$$

The hypotheses can be written as:

$$H_0: p_{1|1} = p_{1|2} = \ldots = p_{1|I}$$

$$H_1: p_{1|1} \leq p_{1|2} \leq \ldots \leq p_{1|I}$$

with at least one strict inequality.

The prediction equation under ordinary least squares fit is:

$$\widehat{p_{1|i}} = x_{1+} + b * (i - \bar{i})$$

where:

$$x_{1+} = \frac{n_{1+}}{n_{++}}, \underline{i} = \frac{\sum_i n_{+i} * i}{n}, and\ b = \frac{\sum_i n_{+i} * \left(x_{1|i} - x_{1+}\right) * (i - \bar{i})}{\sum_i n_{+i} * (i - \bar{i})^2}$$

The Cochran-Armitage test statistic[8,9] is given by:

$$z^2 = \left(\frac{b^2}{x_{1+} * x_{0+}}\right) * \sum_i n_{+i} * (i - \bar{i})^2$$

or: $z = \frac{\sum_i n_{+i} * (x_{1|i} - x_{1+}) * (i - \bar{i})}{\sqrt{x_{1+} * x_{0+} * \sum_i n_{+i} * (i - \bar{i})^2}}$

where: $x_{0+} = 1 - x_{1+}$

Unlike the classical Cochran-Armitage trend test, here we cannot assume that the z-score has an asymptotic normal distribution. Hence, we adopted a permutation-based approach to determine the p-value of significance. We permuted the case-control labels *M* times and calculated z-scores for each permutation. The p-values were calculated as the ratio of the number of times we saw a permuted z-score at least as large as the original z-score, including the original, to the total number of permutations including the original observation.

For the fixed threshold version of our test, we selected variants with gnomAD global minor allele frequencies < 0.05 as qualifying rare variants. For the variable threshold version of our test, we first considered all MAFs observed in the cohort to select an optimal MAF, then we used only the in-cohort MAFs < 0.05 to select an optimal MAF. We performed 10,000 random permutations to generate the p-values in each case. To ensure the validity of our permutation tests, we tried different MAF cutoffs chosen uniformly at random from the range of minimum and maximum MAFs in the original data during each iteration.

As a sanity check, we also applied the combine and collapse (CMC) strategy to collapse the variants at gnomAD global minor allele frequency (MAF) cutoffs of 0.05 in our pathway of interest. Then, we used the one-tailed Fisher's exact test to determine the significance of the difference in burden between PF and sepsis cohorts. A separate analysis was done for each of the following categories of variants: ΔP, neutral missense, and synonymous. To mitigate potential bias due to imperfect ethnicity matching between cases and controls, the burden analysis was performed using only the quality-filtered European PF cases (N=24) and sepsis controls. Gene-based rare variant collapsing analysis was performed on complement system genes using CMC-Fisher[10], KBAC[11], SKAT[12], and VT-Price[13] tests using the RVTESTS package[14] and the KBAC package (https://github.com/gaow/kbac/).

**SOFA score calculation and multivariate logistic regression**

The Sequential Organ Failure Assessment (SOFA) score[15] was calculated for each patient in the study based on review of the medical record (Boston PF Cohort) or on data available through dbGaP (Control Cohort). Scores were not computed for patients missing one or more parameters. Modeling was performed using Graphpad Prism version 9.3.

**Vectors**

LeGO-eGFP-P2A-MSC and LeGO-mCherry-P2A-MSC expression vectors were generated by GenScript Biotech (Piscataway, NJ, USA) as follows. The LeGO-iG2 expression vector (Addgene #27341) was digested with Not1 and BsrG1 to remove the IRES-eGFP cassette and, in its place, a chemically synthesized cassette with a 5' Not1 site, a Kozak sequence, the coding sequence of a fluorescent protein (eGFP or mCherry), a P2A site, a multiple cloning site with Xho1, Xba1, and BstB1 restriction sites, and a 3' BsrG1 site was inserted. Human wild-type cDNA sequences for *ITGAM*, *ITGAX* and *ITGB2* were downloaded from Ensembl Genome Browser. Each wild-type and mutant human *ITGAM*, *ITGAX* and *ITGB2* cDNA sequence was codon-optimized to remove any internal Xho1 and BsrG1 sites, cDNA constructs for each were chemically synthesized with flanking 5' Xho1 and 3' BsrG1 sites, and the cDNA constructs were cloned into LeGO-mCherry-P2A-MSC (in the case of ITGAM and ITGAX constructs) and LeGO-eGFP-P2A-MSC (in the case of ITGB2 constructs). All of the constructs were confirmed by full plasmid sequencing at the MGH CCIB DNA Core Facility (Massachusetts General Hospital Center for Computational and Integrative Biology, Boston, MA, USA).

**Generation and culturing of stable cell lines**

HEK-293T cells (ATCC) were maintained in Dulbecco's modified Eagle's medium (DMEM) supplemented with 10% fetal bovine serum, 100 µg/ml streptomycin, and 100 Units/ml penicillin. Cells were grown at 37 °C and 5% $CO_2$. To generate stable cell lines, HEK-293T cells were co-transfected with different combinations of LeGO-eGFP-P2A-cDNA and LeGO-mCherry-P2A-cDNA constructs using lipofectamine transfection (Thermo Fisher Scientific) according to manufacturer's suggested protocol. After transfection, the cells were sorted for GFP and mCherry double-positivity using a FACSAria cell sorter (BD Biosciences, Mountain View, CA, USA) under BL2+ sterile conditions. Populations with equivalent GFP and mCherry fluorescence intensity were selected across the different CR3 and CR4 cell lines to ensure similar expression levels of the transfected constructs. Continued expression of the constructs was confirmed periodically by flow cytometry for GFP and mCherry using an LSRFortessa flow cytometer (BD Biosciences) and flow cytometry data were analyzed using FlowJo software. HEK-293T cells were periodically tested for *Mycoplasma* contamination using the Venor GeM mycoplasma detection kit (Sigma Aldrich; Natick, MA) and remained negative throughout the study.

**Western blot analysis of cell lysates**

Equal numbers of cells of each flow-sorted CR3 and CR4 stable line were collected in 250 µl HEPES buffered saline (HBS) and then lysed with an equal volume of 2% NP-40 in Tris-buffered saline (TBS) containing HALT protease inhibitor cocktail (Thermo Fisher Scientific). Lysates were mixed at 4˚C for 20 minutes and then subjected to centrifugation at 16,000 RCF for 15 minutes. Equivalent amounts of purified lysate for each cell line were then run on denaturing SDS-PAGE gels and analyzed by Western blot for the presence of integrin subunits using antibodies purchased from Cell Signaling Technology (Danvers, MA): integrin $\alpha_M$ (D6X1N), integrin $\alpha_X$ (D3V1E), and integrin $\beta_2$ (D4N5Z).

**Flow cytometry analysis of cell surface expression of CR3 and CR4**

Cell surface CR3 and CR4 expression was assessed by staining cells with an APC-labeled anti-CD18 antibody (BD BioSciences, 551060). Flow cytometry was performed using an LSRFortessa flow cytometer (BD Biosciences) and data were analyzed using FlowJo software.

**Solid phase iC3b binding assay**

96-well polystyrene plates were coated with iC3b (10 µg/ml) in PBS at 4°C overnight, then blocked with 200 µl 0.05% polyvinylpyrrolidone. Cells (1 x $10^5$ for CR4-expressing cells or 2 x $10^5$ for CR3-expressing cells) in HBS with 1 mM $Ca^{2+}$, 1 mM $Mg^{2+}$, and 0.5 mM $Mn^{2+}$ were added to each well and incubated at 37°C for 20 minutes. The unbound cells were removed by 3 washes with HBS. Adherent cells were stained with 0.5% crystal violet. Excess dye was washed off with distilled water, then the remaining dye was solubilized in methanol and absorbance was measured at 570 nM on a standard plate reader. Data were analyzed using one-way ANOVA with Dunnett's post hoc test after confirming normality of the data with the Shapiro-Wilk test.

**Dual luciferase reporter assay**

96-well plates (Medisorp, Thermo Scientific Cat. #467320) were coated with 1 μg/well of complement protein iC3b (Complement Tech, Cat. #A115) or with mock solution, followed by incubation at 4°C for 6 hours before being washed with PBS. The plates were then blocked with 0.5% polyvinylpyrrolidone in PBS for 1 hour at room temperature and then washed again with PBS. Parental HEK-293T cells and HEK-293T cells expressing wild-type or mutant CR3 or CR4 were seeded in 12-well tissue culture plates at a density of 1 x $10^6$ cells per well. Once cells reached 80% confluency, they were transfected with 75 ng of the pSI-Check2-hRluc-NFκB-firefly plasmid (Promega) using Lipofectamine 2000 (Invitrogen, Cat. #11668-030) and incubated for 6 hours. Following transfection, the cells were harvested and seeded onto the iC3b-coated or mock-coated 96-well plates at 5 x $10^4$ cells per well and were incubated at 37 °C for 18 hours. The cells were then stimulated with 10 ng/ml of TNF-α (Sigma, Cat. #654205-10UG) at 37 °C for 4 hours. The luciferase activity of the cells in response to TNF-α treatment was measured using the Dual-Luciferase Reporter Assay System (Promega, Cat. #E2920) following the manufacturer's instructions.

**Data and code availability**

Sequence variants passing GATK filters that support the findings of this study will be available through dbGaP (currently available upon request from the authors). For statistical analysis we used R v4.0.1. The code for RVTT is available through https://github.com/snz20/RVTT. Source data for all figures are provided with the paper.

**Acknowledgements:** The authors wish to express their gratitude to the patients who participated in this study and their families, as well as to Drs. George Cannelos, Robert Mayer, and Ann LaCasce of the Dana Farber Cancer Institute for their support. The authors also thank Dr. James Knight for his assistance with bioinformatics and members of the Flaumenhaft, Bendapudi, and Losman labs for technical assistance. P.K.B. was supported by Beth Israel Deaconess Medical Center and the National Institutes of Health (R21 HL153655 and K08 HL136840). S.N. gratefully acknowledges support from the Australian Parkinson's Mission through funding from the Australian government. J.A.L. was supported by the Dana Farber Cancer Institute.

**Author contributions:** P.K.B. and J.A.L. conceived of the project, analyzed the data, and assembled and wrote the manuscript; S.S. and S.N. assisted in drafting the manuscript; P.K.B., J.A.L, R.M., W.D., S.R., and S.S. designed experiments and analyzed data; P.K.B. and J.R. developed and conducted the *in vitro* assays and analyzed the data; B.R. generated the stable cell lines; A.R., L.T., and L.C. conducted *in vitro* assays and analyzed data; S.N., O.S., A.T.P., and M.B. performed sequencing quality control and statistical analyses overseen by S.S.; P.K.B., J.R., M.C., B.P., J.K., E.F., J.C.H., O.P., and V.N. developed and operationalized the specimen collection system; all of the authors read and helped to edit the manuscript.

**Competing Interests:** J.-A. Losman has received funding support unrelated to this project via DFCI from Lilly and via the Broad Institute from Bayer.
